# Prospective Assessment of Bone Marrow Involvement with Positron Emission Tomography vs Bone Marrow Biopsy in Patients with Lymphoma

**DOI:** 10.1101/2023.03.21.23287535

**Authors:** Suhas Singla, Sandeep Batra, Pankaj Dougall, Nitin Dayal, Rahul Naithani

## Abstract

**Background:** Bone marrow involvement (BM involvement) in lymphoma is a known adverse prognostic factor. Bone marrow biopsy (BMB) is the gold standard for detection of bone marrow involvement but is invasive modality. Positron Emission Tomography with Computed Tomography (PET-CT) scan has the ability to assess bone marrow involvement. We aimed to assess the concordance of PET-CT for BM involvement with BMB.

**Methods:** 75 consecutive newly diagnosed cases of lymphoma were enrolled and were assessed for BM involvement with PET-CT and BMB.

**Results:** Of 75 patients, eighteen patients (24%) had BM involvement, with 19% (n=14) detected with BMB, and 13 (17%) with FDG 18 PET CT. There was a concordance rate of 88% amongst PET-CT and BMB. It was 92% in Hodgkin’s lymphoma (HL), 71.4% in non-Hodgkin’s lymphoma (NHL), and 91% and 70% in high-grade (HG) and low-grade (LG) NHL, respectively. Sensitivity, specificity, PPV, NPV, and accuracy of PET-CT for study population were 69.23%, 93.44%, 69.23%, 93.44%, and 89.19 % respectively. In patients with NHL sensitivity, specificity, NPV, PPV, and accuracy of PET-CT were 54.55%, 94.23%, 66.67%, 90.74%, and 87.30%, respectively; whereas in HL group these were 100%, 88.89%, 75%, 100%, and 91.67%, respectively.

**Conclusions:** PET-CT has got a high concordance with bone marrow biopsy in detecting bone marrow involvement with high specificity, NPV and accuracy. A high sensitivity, specificity, NPV and accuracy for detecting bone marrow involvement in patients with HL, aggressive B cell NHL, and T cell NHL was observed but the same parameters were not at par in patient with indolent (low-grade) NHL.

## Introduction

Bone marrow involvement varies amongst the various histopathological types of lymphoma and is usually diagnosed with a bone marrow biopsy, an age old gold standard. It is, however, associated with certain disadvantages such as pain, invasiveness, hemorrhagic manifestations, and inability to inform about the marrow as a whole and missing certain focal lesions.^1^,^2^,^3^ PET-CT, is an imaging modality widely used in staging of lymphoma and can demonstrate BM involvement with high sensitivity and specificity. However, in certain conditions like infection, inflammation, and anemia it can be falsely positive.^4^ There are larger prospective and retrospective studies available. However, prospective and even retrospective data from India is scarce.

In this study, we prospectively tried to find the concordance between PET-CT and bone marrow biopsy for BM involvement in patients with lymphoma and its subtypes.

## Patients & Methods

Newly diagnosed patients of lymphoma with age ≥18 years were enrolled prospectively. Patients with recurrent, relapsed cases of lymphoma, or with any other concurrent malignancy and who were ineligible to undergo PET-CT or bone marrow biopsy were excluded.

Bone marrow biopsy was interpreted by the expert hematopathologist morphologically and with immunohistochemistry. A positive bone marrow involvement result was defined by focally increased FDG uptake or diffusely increased bone marrow uptake with an intensity greater than that of the liver as assessed by the nuclear medicine expert in the bone marrow

### Statistical Analysis

Data were expressed in terms of percentage and means with standard deviation and median with interquartile range. Fisher’s and chi-square test, were used for categorical variables; student t test was used for continuous variables. In all analyses, P <0.05 was considered statistically significant. Sensitivity, specificity, positive predictive value (PPV), negative predictive value (NPV) and accuracy with Clopper–Pearson exact confidence limits were calculated.

## Results

A total 75 patients fulfilling the inclusion and exclusion criteria were enrolled presenting from Sep 2018 to Jan 2020. The baseline clinical characteristics are demonstrated in table I. Diffuse large B cell lymphoma was the most common pathology (52%). Most patients presented with stage IV disease (43%) and had B symptoms (59%).

**Table I:** Demographic profile of study population.

|  |  |
| --- | --- |
| Characteristics |  |
| Age | 56.4 ± 17 years |
| Sex |  |
| Male | 50 (67%) |
| Female | 25 (33%) |
| Subtypes |  |
| Diffuse Large B Cell Lymphoma | 39 (52%) |
| Hodgkin's Lymphoma | 12 (16%) |
| T Cell Lymphoma | 9 (12%) |
| Follicular Lymphoma | 7 (10%) |
| Marginal Zone Lymphoma | 3 (4%) |
| Primary CNS Lymphoma | 2 (3%) |
| Burkitt Lymphoma | 1 (1%) |
| Mantle cell Lymphoma | 1 (1%) |
| Primary mediastinal B Cell Lymphoma | 1 (1%) |
| Stage |  |
| Stage I | 15 (20%) |
| Stage II | 9 (12%) |
| Stage III | 19 (25%) |
| Stage IV | 32 (43%) |
| ECOG performance status |  |
| "0" to "1" | 46 (61%) |
| "2" to "4" | 29 (39%) |
| B symptoms | 44 (59%) |

Eighteen (24%) patients had BM involvement, and there was no statistical difference in the baseline demographic, clinical, hematological, serological, and pathological parameters in patients with and without BM involvement except for difference in Ki-67 (marker of proliferation); 41.9 ± 28.1 vs 66.3 ±24.7% (p-value-.001).

BMB and PET-CT detected BM involvement in 14 (19%) and 13 (17%), respectively. Considering BMB as gold standard, the sensitivity, specificity, positive predictive value (PPV), negative predictive value (NPV), and accuracy of PET-CT were sensitivity 69.2, 93.4%, 69.2%, 93.4%, and 89.1%, respectively. PET-CT was concordant with bone marrow biopsy in 88% cases and detected 4 (5%) additional patients with BM involvement and missed 5 patients as detected by bone marrow biopsy. However, on independent analysis for subtypes, maximum concordance (100%) was observed in T cell NHL amongst patients with NHL followed by 92% each in DLBCL and HL. Least concordance was observed in LG-NHL (70%). Statistical analysis for sensitivity, specificity, PPV, NPV and accuracy were as shown in table II where the sensitivity was lowest in HG-NHL (50%) and highest in HL and T cell NHL. Specificity and accuracy were lowest in LG-NHL. 100% NPV was observed in T cell NHL and HL. Impact of various prognostic factors on the concordance, sensitivity, specificity, PPV, NPV, and accuracy of PET-CT is shown in table II.

**Table II:** Concordance rate of BM and PET CT as per lymphoma subtype.

| Type |  |  | BMB |  | Total | p-value | Concordance | Sensitivity (%) | Specificity | PPV (%) | NPV (%) | Accuracy (%) |
| --- | --- | --- | --- | --- | --- | --- | --- | --- | --- | --- | --- | --- |
|  |  |  | No | Yes |  |  |  | (CI 95%) | (CI 95%) | (CI 95%) | (CI 95%) | (CI 95%) |
| Overall | PET-CT | No | 57 | 5 | 62 | <0.001 | 88 | 64.29 | 93.44 | 69.23 | 91.94 | 88.00 |
|  |  | Yes | 4 | 9 | 13 |  |  | 35.14- 87.24 | 84.05- 98.18 | 44.68-86.24 | 84.91-95.85 | 78.44- 94.36 |
| HL | PET-CT | No | 8 | 0 | 8 | 0.018 | 92 | 100 | 88.89 | 75.00 | 100 | 91.67 |
|  |  | Yes | 1 | 3 | 4 |  |  | (63.06- 100.00) | (51.75-99.72) | (32.10-95.01) |  | (61.52 - 99.79) |
| NHL | PET-CT | No | 49 | 5 | 54 | <0.001 | 93 | 54.55 | 94.23 | 66.67 | 90.74 | 87.3 |
|  |  | Yes | 3 | 6 | 9 |  |  | (23.38 - 83.25) | (84.05 - 98.79) | (37.04 - 87.18) | (83.64 - 94.95) | (76.50 - 94.35) |
| LG-NHL | PET-CT | No | 4 | 2 | 6 | 0.524 | 70 | 60 | 80 | 75 | 66.67 | 70 |
|  |  | Yes | 1 | 3 | 4 |  |  | (14.66- 94.73) | (28.36-99.49) | (31.11- 95.22) | (38.55- 86.44) | (34.75- 93.33) |
| HG-NHL | PET-CT | No | 45 | 3 | 48 | 0.008 | 91 | 50 | 95.74 | 60 | 93.75 | 90.57 |
|  |  | Yes | 2 | 3 | 5 |  |  | (11.81-88.19) | (85.46- 99.48) | (23.70- 87.87) | (87.05-97.10) | (79.34-96.87) |
| DLBCL | PET-CT | No | 34 | 1 | 35 | 0.023 | 92 | 66.67 | 94.44 | 50 | 97.14 | 92.31 |
|  |  | Yes | 2 | 2 | 4 |  |  | (9.43-99.16) | (81.34-99.32) | (17.27-82.73) | (87.26-99.14) | (79.13-98.38) |
| T NHL | PET-CT | No | 7 | 0 | 7 | 0.028 | 100 | 100 | 100 | 100 | 100 | 100 |
|  |  | Yes | 0 | 2 | 2 |  |  | (15.8-100) | (59.04-100) |  |  | (66.37-100) |

HL-Hodgkin lymphoma; NHL-Non-hodgkin lymphoma; LG-NHL-Low grade Non-hodgkin lymphoma; HG-NHL-High grade Non-hodgkin lymphoma; DLBCL-Diffuse large B cell lymphoma; T-NHL-T cell non-hodgkin lymphoma

## Discussion

Determination of the presence of BM involvement is an important factor as its presence upstages the disease, portends more chances of cytopenia with therapy and may yield to poor stem cell yield. Bone marrow is an invasive test and any non-invasive modality which could replace it is desirable. PET-CT has been explored in various retrospective and prospective studies for its potential to detect BM involvement as shown in table III. However, it had not been very well explored in the Indian subcontinent with a few studies available to mention. We assessed the impact of various prognostic factors on the effectivity of PET-CT to determine BM involvement. PET-CT has high concordance of 78-92% with bone marrow biopsy in detecting BM involvement as depicted in table 5. and was well in coherence with our study where we found it to be 88%. Our findings of sensitivity, specificity, PPV, NVP and accuracy were similar to the data from the previous studies. BM involvement varies amongst the various types of lymphoma varying from <1% to >60% in HL and 10% to >90% in NHL.^5^ When analyzed for various subtypes, PET-CT had a 92% concordance and 100% sensitivity in patients with HL, a specificity of 88.89% as one patient detected as having BM involvement on PET-CT was considered as false positive for statistical analysis. Previous studies showed similar findings for predictive and accuracy values. PET-CT is known to have a high a sensitivity, specificity, PPV, NPV and accuracy in patients with HL, with a potential to detect additional patients with BM involvement. In our study PET-CT picked-up all the patients with BM involvement and detected 1 additional patient. Adams et al. in a metanalysis reinforced the same and concluded that BMB can be replaced with PET-CT in a newly diagnosed case of HL.^6^ In patients with NHL, PET-CT tends to have a lower sensitivity and specificity than in patients with HL as found by Pakos et al in the landmark metanalysis.^7^ Though, sensitivity was lower in our study too, we found a higher specificity which may be because of higher number of patients with HG-NHL in this subgroup. PET-CT tends to have a higher concordance, sensitivity and accuracy in patients with HG-NHL as was found in another metanalysis by Chen et al.^8^ Further, in patients with T cell NHL, all the statistical values amounted to 100%, signifying a high rate of concordance as had been previously found in few studies analyzing PET-CT in detection of BM involvement T cell NHL patients.^9^ PET-CT has got a high NPV in most of the subtypes of lymphoma except LG-NHL, thus can be utilized for avoiding a painful and invasive procedure like BMB in newly diagnosed case of HL and HG-NHL. We tried to look for certain factors which can determine a subset of patients where BMB can be avoided as PET-CT tends to have a high sensitivity and NPV in those patients. On assessing for the effect of ECOG performance status (ECOG-PS) (“0 to 1” vs “2 to 4”), PET-CT had similar concordance and a higher sensitivity, NPV and PPV in ECOG-PS “2 to 4” subgroup. Similarly, in patients with B symptoms and high serum lactate dehydrogenase PET-CT had a higher sensitivity but no gross difference in NPV was found. In patients with HL, prediction rule for BM involvement by Vassilakopoulos et al. can be used to avoid any theoretical risk of missing BM involvement. BM involvement in patients with LG-NHL should always be ruled out with an iliac crest biopsy.^1^

**Table III:** Summary of prospective studies.

| Author | Year | Number of patients | Country of origin | Type of lymphoma | Concordance | Sensitivity | Specificity | PPV | NPV | Accuracy |
| --- | --- | --- | --- | --- | --- | --- | --- | --- | --- | --- |
| Prospective (Indian) |  |  |  |  |  |  |  |  |  |  |
| Current study | 2020 | 75 | India | Overall | 88 | 69.2 | 93.4 | 69.2 | 93.4 | 89.2 |
|  |  |  |  | HL (n=12) | 92 | 100 | 88.9 | 75 | 100 | 91.7 |
|  |  |  |  | NHL (n=63) | 71.4 | 54.6 | 94.2 | 66.7 | 90.7 | 87.3 |
| Mittal et al<sup>10</sup> | 2011 | 97 | India | Overall | 92 | 89 | 93 | 89 | 93 | NA |
|  |  |  |  | HL (n=20) | 90 | 100 | 86 | 71 | 100 | NA |
|  |  |  |  | NHL (n=77) | 92 | 100 | 94 | 93 | 100 | NA |
| Prospective (International) |  |  |  |  |  |  |  |  |  |  |
| Xiao-Xue et al<sup>11</sup> | 2020 | 153 | China | Overall | 74.5 | 54.3 | 80.5 | NA | NA | 74.5 |
|  |  |  |  | HL (n=15) | 47 | 0 | 46.7 | NA | NA | 46.7 |
|  |  |  |  | B-NHL (n= 59) | 88.1 | 80 | 90 | NA | NA | 88.1 |
|  |  |  |  | FL (n = 24) | 71 | 37.5 | 87.5 | NA | NA | 70.8 |
|  |  |  |  | T-NHL (n = 34) | 76.5 | 57.1 | 81.5 | NA | NA | 76.5 |
| Voltin et al<sup>12</sup> | 2018 | 832 | Germany | HL | 87 | 95 | 86.5 | 14.7 | 99.9 | NA |
| Özpolat et al<sup>13</sup> | 2018 | 38 | Turkey | Overall | 57 | 77 | 51 | 88 | 33 | NA |
|  |  |  |  | NHL (22) | 68 | NA | NA | 57 | 95 | NA |
|  |  |  |  | HL (16) | 37.5 | NA | NA | 90 | 100 | NA |
| Alzahrani et al<sup>14</sup> | 2016 | 530 | Canada | DLBCL | 76 | 60 | 79 | 36 | 91 | NA |
| Corte´s-Romera et al<sup>15</sup> | 2014 | 147 | Spain | DLBCL/ HL | 87 | 95 | 86 | 87 | 54 | 99 |
| Shih-Feng Cho et al<sup>16</sup> | 2015 | 185 | China | Overall | 82.1 | 50 | 91.6 | 63.6 | 86.2 | NA |
|  |  |  |  | aB-NHL | 88.1 | 70 | 92.1 | 66.7 | 93.2 | 88 |
|  |  |  |  | HL | 93.8 | NA | NA | NA | 100 | NA |
|  |  |  |  | FL & MZL | 65-67 | 27.8 | 89.7 | 62.5 | 66.7 | 65.9 |
|  |  |  |  | T-cell | 76.9 | 50 | 88.9 | 66.7 | 80 | 76.9 |
| El-Galaly et al<sup>17</sup> | 2012 | 454 | Germany | HL | 85 | 85 | 86 | 28 | 99 | 86 |
| Ngeow et al<sup>18</sup> | 2009 | 122 | Singapore | B-NHL (n=63) | 90 | 41 | 95 | 91 | 58 | NA |
|  |  |  |  | B-NHL (n=21) | 67 | 57 | 95 | 57 | 95 | NA |
|  |  |  |  | T cell NHL (n=17) | 100 | 100 | 100 | 100 | 100 | NA |
|  |  |  |  | HL (n=21) | 90 | 100 | 90 | 33 | 100 | NA |
| Carr et al<sup>19</sup> | 1998 | 50 | UK | NHL (38) /HL (12) | 78 | NA | NA | NA | NA | NA |
| Moog et al<sup>20</sup> | 1998 | 78 | Germany | NHL (39)/HL (39) | 82.1 | 81 | 100 | NA | NA | NA |
| Retrospective (Indian) |  |  |  |  |  |  |  |  |  |  |
| Agrawal et al<sup>21</sup> | 2013 | 38 | India | HL (paediatric) | 81.6 | 87.5 | 100 | 100 | 96 | NA |

The limitation of our study was that no quantitative measures such as SUV was used and only qualitative assessment PET-CT was done. No PET-CT directed biopsy were performed which may have increased the yield, though most of the sites were not accessible or the patients refused. Sample size was small, but yet adds information regarding the role of PET CT in assessing the bone marrow.

## Conclusion

PET-CT has got a high concordance with bone marrow biopsy in detecting bone marrow involvement with high specificity, NPV and accuracy. It has got a high sensitivity, specificity, NPV and accuracy for detecting bone marrow involvement in patients with Hodgkin’s lymphoma, aggressive B cell non-Hodgkin’s lymphoma, and T cell non-Hodgkin’s lymphoma.

## Data Availability

All data produced in the present work are contained in the manuscript

